# Nurses’ experience of using video consultation in a digital care setting and its impact on their workflow and communication

**DOI:** 10.1101/2022.02.23.22271275

**Authors:** SeyedehMaryam Razavi, Nasim Farrokhnia, Nadia Davoody

## Abstract

Sweden as many other countries uses video consultation to increase patients’ access to primary healthcare services particularly during the COVID-19 pandemic. Working in digital care settings and using new technologies, in this case video consultations, require learning new skills and adoption to new workflow. The aim of this study is to explore nurses’ experience of using video consultation in a digital care setting and its impact on their workflow and communication. Fifteen semi-structured interviews were carried out with registered nurses recruited from a private digital healthcare provider. Interviews were recorded, transcribed, and analysed using an abductive approach. Nurses’ workflow was modeled, and several categories and subcategories were identified: nurses’ workflow (efficiency, flexibility, and information accessibility); communication (interaction with patients and interprofessional communication); user experience (change and development of the platform, challenges, and combining digital and physical care). Even though providing online care has its limitations, the nurses were positive towards using video consultations.

## Introduction

The complex nature of healthcare, an aging population suffering from multiple chronic diseases, and occurrence of the COVID-19 pandemic require new ways of providing care to patients and citizens. The traditional ways of providing care to all patients are not sufficient due to lack of resources [1]. Information and communication technology has been used widely in healthcare, and telemedicine has been seen as one of the essential solutions to this matter [2]. E-health innovations enable patients and citizens to among others obtain healthcare online, get involved in their own care, and track their disease and treatment processes.

In recent years, receiving care through video consultations has been an inseparable part in health care. Video consultations, digital care visits, video visits, and video consultations are some of the terms for receiving care online [3–6]. Video consultation is defined as a direct and synchronous conversation between healthcare professionals and patients [7]. With video consultations in this study, we mean real-time tele consultation/virtual visit that is initiated by a patient at a distance using information and communication technology. New paragraph: use this style when you need to begin a new paragraph.

### The use of video consultations in Sweden and globally

Currently, the use of video consultations has increased significantly worldwide due to the occurrence of the COVID-19. Various countries, including the United States, UK, Japan, China, and several European countries such as Sweden, plan and develop healthcare strategies based on digital services. In this model, video counseling services are provided by private companies to patients to deliver primary care services [8]. With the onset of the COVID-19 pandemic outbreak in March 2020, the use of telemedicine increased to reduce the risk of spreading the infection, especially in countries such as the USA, UK, China, and Australia. In recent years, particularly during the COVID-19 pandemic, several studies have been performed focusing on the use of video consultations and telemedicine, its benefits, and challenges in health care from healthcare professionals’ and patients’ perspective [9–19].

In Sweden, video consultations have been provided by private healthcare providers to citizens and patients since 2016. [20] Patient satisfaction with digital care provided by private healthcare providers such as KRY AB and MinDoktor have been around 90 percent [21]. Currently, most of the video consultations are done by these two private, for-profit companies established in 2014 and 2015. In Sweden, digital care users have mostly been from large regions such as Stockholm region, Västra Götaland county, and region of Scania and most common diagnoses have been infections, skin conditions, intestinal conditions, and male’s or female’s specific health problems [21].

### Socio-technical model

Designing health information systems and eHealth services for providing care online requires having a socio-technical approach in mind as they affect the work processes of care professionals and the way they provide care to the patients and citizens. The success of a health information system is influenced by technical, social, organizational, and socio-political factors that ultimately affect the patient’s outcome in healthcare systems. In this regard, one of the models employed for the comprehensive analysis of health information systems is the social-technological model. Hardware and software computing infrastructure, clinical content, human computer interface, people, workflow and communication, internal organizational features, external rules and regulations and measurement and monitoring are the 8 dimensions of the socio-technical conceptual model [22]. Profound study and understanding of these eight dimensions will assist us to ensure that every patient receives the right care at the right time [23]. Workflow and communication as one of the dimensions of the socio-technical model acknowledge the collaboration of the healthcare team to accomplish patient care.

Usually, new eHealth solutions do not comply with the actual “clinical” workflow. In such a case, modifying the workflow or changing the health information technologies need to be considered. Numerous studies have been done regarding the impact of technology on nurses’ workflow and Communication [24–33]. Research on the impact of technology on nurses’ workflows in the physical settings shows that technology has a positive impact on nurses’ workflows. Having access to information at the point of care, improved collaboration, job performance and communication, improved patient safety and reduction of medication errors, and improved nurses’ perception of the system’s role in their workflow have been some of the benefits of using technology in a physical setting. The studies have, in addition, addressed some challenges such as technical problems, difficulties in providing clinical assessments, clinicians’ skepticism and criticism, disconnection with the IT team, and setting up and maintenance of the devices [33–37]. In digital settings, however, the impact of video consultations on nurses’ work process and communication has not been studied in detail. To support nurses in a digital care setting we need to obtain a deep understanding of their workflow and the impact of the new technology, in this case video consultation, on their workflow and communication.

### Aim

The aim of this study is to explore the nurses’ experience of using video consultations in a digital care setting. In addition, the aim is to study the opportunities and limitations with video consultations and its impact on the nurses’ workflow and communication in Sweden. Studying these aspects will assist us to support nurses throughout their work processes and also improve the design and implementation of these services in future.

## Materials and methods

Qualitative research methods were used, and data was gathered through semi-structured interviews with registered nurses at KRY AB in Sweden. The socio-technical model was used to develop the interview guide in this study and our focus has been on the workflow and communication of nurses in a digital setting.

### Data collection

Fifteen interviews were performed in this study. Each interview took about 30 minutes and was conducted by telephonic or video calls. Recruitment of the participants was done in the beginning of 2020. All interviews were audio recorded and transcribed verbatim. Interviews were analyzed using an abductive approach [38]. Questions asked of individuals were regarding their workflow and communication.

### Participant selection and setting

In this study, we involved nurses working in one of the private digital care providers, namely, KRY AB as they are one of the first digital care providers in Sweden. KRY AB has in 2018 started their physical clinics to provide care even physically if needed.

These integrated digital and physical services are called Digi-physical services and are aimed to close the gap between digital and physical care and facilitate care flow both for patients and healthcare workers [39–41]. The inclusion criteria were specified as follows: English speaking participants with at least six months experience of working in physical and digital care settings. All the participants were registered nurses and had experience of working in physical settings with a mean average of 17 years (range 2-41). All participants have been working at least 6 months with the digital platform at KRY AB. Two females and 13 males were interviewed in this study. Table 1 indicates the characteristics of the participants.

**Table 1.** Participant characteristics.

| Age | Years of experience in<br>physical setting | Years of experience in<br>digital care setting |
| --- | --- | --- |
| 30-39 | 13 y | 1 y |
| 30-39 | 17 y | 1y |
| 40-49 | 20 y | 1y |
| 40-49 | 26 y | 1y |
| 60-69 | 35 y | 1y |
| 40-49 | 25 y | 1y |
| 40-49 | 17 y | 1y |
| 40-49 | 11 y | 7 m |
| 40-49 | 10 y | 1 y, 6 m |
| 40-49 | 24 y | 1 y, 6 m |
| 50-59 | 41 y | 2 y |
| 40-49 | 5y, 6 m | 6 m |
| 40-49 | 2 y | 6 m |
| 40-49 | 14 y | 1 y |
| 20-29 | 3 y | 1 y |

### Data analysis

All three authors (MR, NF, and ND) were involved in creating a preliminary semi-structured interview guide. The interview guide was divided into three different parts focusing on the workflow, communication, and user experience of nurses in the digital setting. The themes were used as key concepts to guide the coding process and the codes under these themes come from analysis of the interview material. All interviews were conducted by the first author (MR) and the preliminary data analysis was performed by her. The preliminary codes were discussed with the co-authors (ND and NF). All authors were involved in the discussion of final codes and categories in this study.

Based on the interviews, the nurses’ workflow was modeled by Camunda, a desktop application for modeling BPMN (Business Process Modeling Notation) [42] workflows.

## Results

The results will be presented in two parts. The first part includes nurses’ workflow model followed by a presentation of the themes, categories, and sub-categories.

### Nurses’ workflow model

To have an overview of how nurses work in a digital setting, their work process has been modeled in this study (figure 1). The process starts by the nurse logging in to the communication platform (Slack) and KRY platform. The technical issues are checked to ensure proper audio, video, and Internet performance. If any technical problems occur, the nurse either tries to solve the problem by herself/himself or contact the technical support at the company. The work process will continue as soon as the problem is solved. In the next step, the nurse sees a list of patients with their names and pictures on their user page. In this section, the nurse chooses a patient that popped up on the screen.

**Figure 1.**
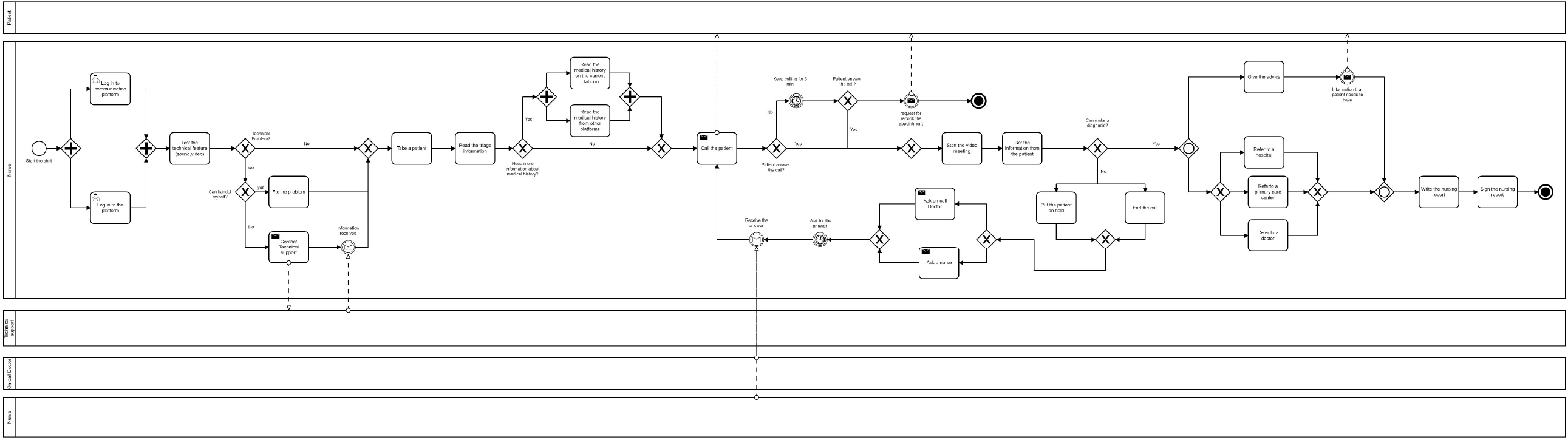
Nurses’ work process illustration.

In the next step, the nurse can study the patient’s chief complaint. It is important to note that when a patient books an appointment, the patient should answer triage questions that indicate what the patient’s main problem is. When the nurse reads the patient’s responses to triage questions, he or she may need to know more about the patient’s previous medical history. The nurse can either choose to read the patient’s previous medical record on the KRY platform or read the patient’s medical record from other platforms where the patient’s information is located, for example, national patient overview (NPÖ). The national patient overview is a web-based tool that gives authorized healthcare professionals access to an overview of a patient’s medical record at other healthcare providers [43]. The next step is calling the patient through the KRY platform. At this stage, if the patient does not respond, the nurse will continue calling her/him for another three minutes, and then sends a text message to the patient. In this message the patient is asked to book another appointment which will end the process. If the patient answers the call, a video meeting will be started by the nurse.

To be able to make a diagnosis, the nurse obtains necessary information from the patient by asking questions related to the patient’s problem. If the nurse is unable to make a diagnosis, she/he can either put the patient on hold or end the call and contact the patient after consulting a doctor or nurse. After consultation with the on-call doctor/nurse, the nurse receives an answer related to diagnosis and should call the patient again. If the nurse doesn’t need any help from the on-call doctor/nurse, she/he can provide a diagnosis directly and continue the process.

The nurse then gives advice and/or referral to a hospital, a primary care center, or a physician. Also, at this stage, the nurse can send a message containing the information that was provided during the meeting to the patient. The nurse writes then a report and documents all the information and interventions. The nurse approves and signs the report which ends the work process.

All participants in this study were working both digitally and physically at different healthcare centers. The results including themes, categories, and subcategories will be presented below (Figure 2).

**Figure 2.**
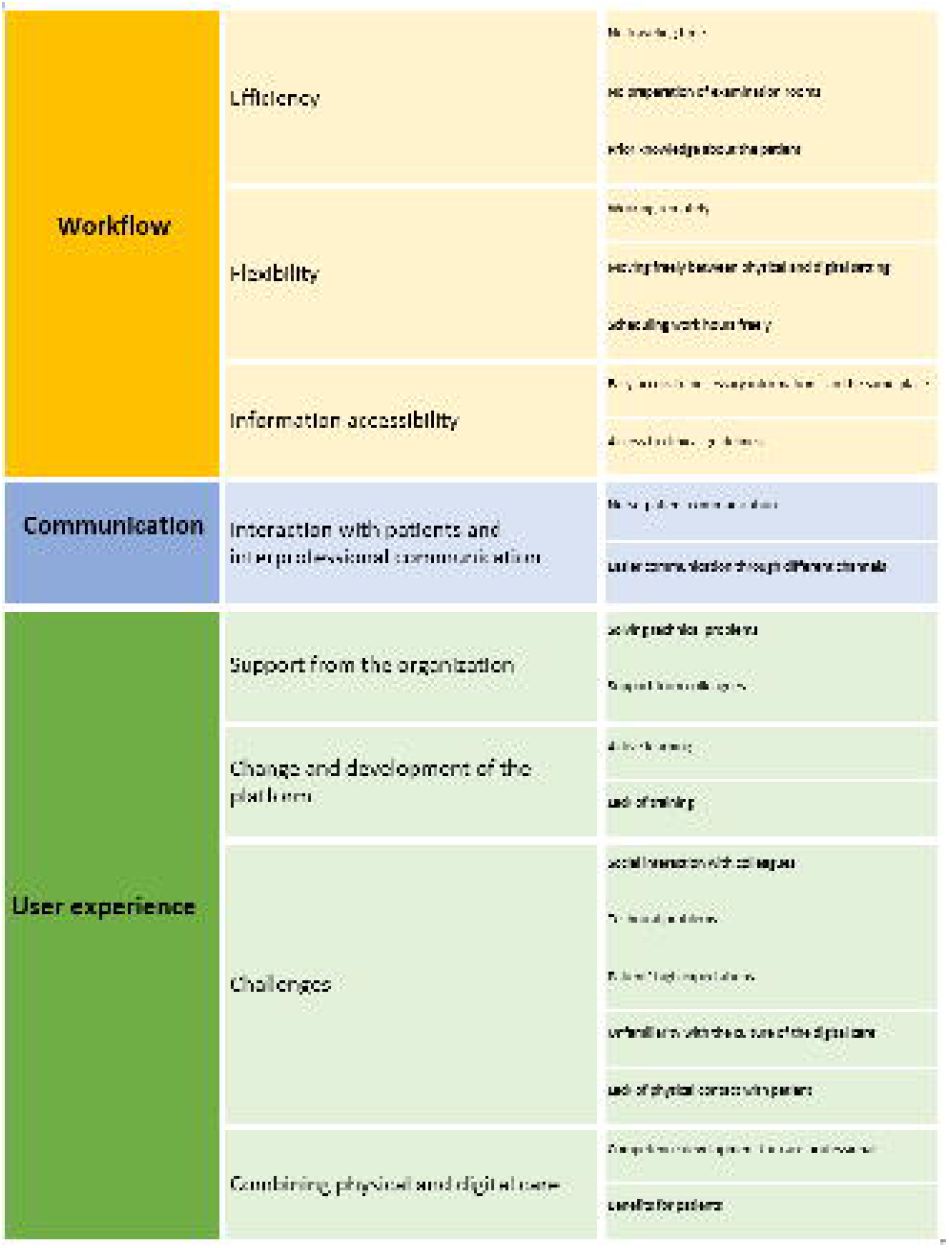
An overview of identified themes, categories, and subcategories.

### Theme 1. Workflow

#### Efficiency

Participants were interested in working with maximum productivity and minimum time or energy wasting e.g., not spending time traveling to the workplace, meeting the patients with short preparation time, and having enough information about the patient. *No travelling time*. Most of the participants stated that it is positive to not spend the time commuting between workplace and home.

*‘I don’t have to spend time traveling to work. Don’t have to spend time traveling home. I have a setting at home which is very comfortable, which is nice at covid-19 time*.

*Working from home.’ (Interview 1)*

##### Not preparing the examination rooms

They also mentioned that not preparing the examination room resulted in having more time for patients. They felt that they were more concentrated in communication with the patient through video consultations than the physical meetings.

*‘It is a lot faster. There is less time spent looking for the patient, preparing the patient, going into the room, sitting down, washing your hands, everything like that. You just kind of start and it is all there.’ (Interview 1)*

##### Prior knowledge about the patient

Most of the participants showed an interest in having as much as possible information about a patient’s medical history in advance.

This helped them to be prepared during the video consultation. They were also satisfied with the information that they got before the visit via NPÖ.

*‘The flow with the patients is much better because before I give them a call, I can read what they are seeking in the form. I can also check NPÖ to check some background information if I do feel I need to. And then when I have all this stuff, I call them. I think it is easier.’ (Interview 3)*

*‘You get the patient’s personal number, and you can read the symptoms and you can see the medical record and read about the medical history and then join the meeting…’* (Interview 13)

Access to the patient’s triage questions and the patient’s health data make the nurses’ workflow and communication more effective.

*‘Before I give them a call, I can read what they are seeking in the form. Also, I can check NPÖ to check some background information. I am much more effective because I have all the facts before the patient calls, mostly I have a quite good picture of what is waiting for me, because of that it is much more effective.’* (Interview 3)

#### Flexibility

Most participants were positive towards having freedom to choose the time and place to work. They also liked the way they could schedule their working days and being able to choose freely to work in a physical or a digital setting.

##### Working remotely

Most of the participants were satisfied working from home.

*‘I am satisfied because I do love this way of working. I can work from home. I can sit on the sofa if I only have this white background.’* (Interview 3)

*‘You don’t have to go to the healthcare center, you just need to talk to someone that tells you what to do. I can do my job from my home.’* (Interview 5)

However, some of the participants have mentioned that working from home was challenging for them as it reduced their social interactions with other colleagues.

*‘I like working from home. I have other work that I also do from home, so I don’t have to go to my workplace. Sometimes you get bored, because you need to talk to someone, but we have a good chat channel [Slack] at KRY, we can talk to each other…’* (Interview 6)

##### Moving freely between physical and digital setting

Participants stated that working with video consultation allows them to work in the physical setting as well. Participants stated that in this way they can practice their skills and be in touch with the patients physically.

*‘… For me also I can work in a physical setting too, but it is up to me. I can change between physical and digital settings.’* (Interview 1)

*‘And all the time with video consultation is hard and if you can combine it then it is good. And also, it is good that patients can meet the nurse or doctor physically… I think in a long time it will be boring with digital and when you are educated you want to do some practice…’* (Interview 13)

*‘I would say if I can choose both, I would choose both 50-50. I love the combination. 100% digitally will be too much. Sometimes I need to see the patient*…*’ (Interview15) Scheduling the working hours freely*. Nurses declared working in a digital setting gives them more autonomy regarding planning their schedule. In physical settings, they need to work 7 hours in predetermined shifts, while in this digital care setting, they can decide about their working hours.

*‘I can decide when I want to work or not. Or I don’t need to work 7 hours in a row. Working or going for one hour or two hours.’* (Interview 2)

#### Information accessibility

Majority of the participants expressed that using the platform is easy because they have access to all the necessary information about the patients. They have access to the KRY medical record as well as NPÖ.

##### Easy access to necessary information is in the same place

The participants were satisfied that they have access to all the information in one place.

*‘It is very easy to follow. Why is it easy to follow? You don’t have many options. You get your patient and patient’s medical record, and you don’t have to write any personal number or other details. Everything is just there…It is easy to follow the previous medical record. Easy to follow and easy to see what it is.’* (Interview 7)

##### Access to clinical guidelines

Participants felt that their diagnosis had a better quality as they could check it with clinical guidelines while talking to the patients. Also, if nurses are in doubt about their diagnosis, they can confirm their diagnosis without any significant delay and simultaneously with a doctor’s approval.

*‘[In a video consultation, the diagnosis is] a lot easier. In the physical setting if you need to check something, you need to talk to the patient and then go to your computer and then talk to the patient which takes time and is not a very professional way. Now I can check things up with quality standards that we do have, what is KRY policy on this and then I check this up. Meanwhile I am talking to the patient, and they aren’t even aware that I’m checking it. It is more quality, but I would say it is effective in that way in the digital care setting.’* (Interview 1)

### Theme 2. Communication

#### Interaction with patients and interprofessional communication

Fast communication between colleagues is more feasible in the digital care setting due to different communication services such as Slack.

##### Nurse – patient communication

Participants were satisfied that nothing disturbs them, so they focus on their work.

*‘There is nothing disturbing me. I have this patient and only this patient, not a colleague knocking on the door, so I can focus on what I do.’* (Interview 3)

*‘Here the only thing that I do is to answer the patient. That is the only thing that I do*…*’* (Interview 6)

##### Easier communication through different channels

Communication between colleagues is more feasible in the digital care setting due to the different communication channels such as Slacks. Several participants indicated that it is much easier to communicate as all the nurses are using Slack, phone or email. Having access to all responses in Slack has made this communication channel to a learning tool.

*‘Slack, it is a learning platform too.’* (Interview 5)

*‘Slack, one thing is good, you can write something to anyone, and they can read it any time they can. I don’t want to interrupt them. Slack is better than physical. In a physical setting two people should be at the same place at the same time and it is hard to get a time to suit both persons. In Slack you write a message and after 10 min you receive a response.’* (Interview 15)

However, some participants stated that communicating in a written form can be problematic as it is more difficult to be understood, get a response in time or receive several responses at the same time.

*‘Negative, it is always complicated when the conversation is in a written form because I hear my words in one way when I am writing them, when the receivers read it, it might sound different for them*…*’* (Interview 15)

### Theme 3. User experience

#### Support from the organization

The majority of the participants mentioned that they receive significant support from the organization regarding technical problems. They also mentioned that they have access to collegial support.

##### Solving technical problems

At the beginning of all working shifts, nurses need to check the technical features including sound and video. If they face any problems, they can contact the technical support via email, phone call or Slack. Participants expressed that they always receive support without any significant waiting time.

‘*There is a channel for this too. They always check the channel. If I write something there, especially a technical problem, they are very fast to fix the problem.’* (Interview 1)

*‘We send messages everywhere and we get quick responses from all of us. So, from someone who knows something. I usually write to them [IT people] in Slack.’* (Interview 5)

##### Support from colleagues

All participants have the possibility to ask their questions through a phone call to the assigned physicians/specialists. They can even send a message to all other doctors/nurses through Slack. In both cases, they will receive an answer without significant waiting time. Unlike the physical setting, nurses do not need to chase the doctor or wait a long time to get a response. According to the participants, the doctors set aside time for them in the digital care setting. This setting allows them to communicate with other colleagues e.g., other nurses, physicians/specialists, and technical experts, easily, if needed.

*‘If I need to know something in general, I can send a picture and write the description as a question and then I will have my answer. Sometimes I can ask a specific doctor. We have a nurses’ group. Also, we can ask each other there. There we can also call but often we write to each other because it is quicker. […] There is a big difference between physical care and digital care. In physical care you always have to chase the doctor and often don’t have time for helping nurses. But [in the digital care setting] they always have time.’* (Interview 4)

*‘If you want to ask something, you have quickly an answer from a doctor or another nurse.’* (Interview 2)

Some participants, however, were more positive towards receiving support from colleagues in a physical care setting.

*‘Also I can mute myself and call a colleague if I don’t get a direct response through Slack. I can put the patient on hold and call a colleague. In a physical setting you have someone inside you can just go and ask a colleague, you don’t have to wait to respond, and you can find the person there and talk to him. It can be a bit negative for digitally. Sometimes in Slack you don’t get a response.’* (Interview 14)

#### Change and development of the platform

The platform was developed regularly, and nurses needed to stay updated all the time.

##### Active learning

The majority of the participants stated that the digital care setting is changing rapidly and that these changes are positive and based on a need. They believed that rapid changes in the platform is practical, and that improvement of the platform does not change the basic structure of their work.

*‘It is changing fast because they are trying to improve it. KRY always changes fast but it is good changing. If they see the change is not good, they put it back. They do not struggle with bad things. It is under construction all the time. It is happening all the time. But I like it.’* (Interview 4)

However, some of the participants mentioned that being updated about all the changes made in the platform is time consuming and they need to put effort to learn about these changes in their free time, without any reimbursement.

*‘I feel in a perfect world they would give us 15 minutes or once a week a time to get through the updates. It is difficult to keep track of all the updated procedures and you should do it in your free time. It is not the way it should work in health care. You should get time to go through the updated information.’ (Interview 8)*

##### Lack of training

Some of the respondents mentioned that the occurrence of all these changes makes it difficult for them to stay up to date when they are away from the digital care setting for a long time. They experienced that they need to have a training course to review all the changes to be able to start working again. In the physical care setting, according to some participants, the changes occur slowly, and it makes it possible for them to be away from their work without the need for training.

*‘…in the KRY platform if you are away for 6 weeks everything has changed. A lot of it has changed anyway. Then you have to kind of restart your training. You can [be away for 6 weeks] in the physical setting because things rarely change that fast. When you come back after 6 weeks or whatever else, maybe they rolled a plastic container to another side of the room.’* (Interview 1)

*‘Sometimes there are a lot of updates with no administration time to read them. If I want to read, I either need to do it in my spare time and I will not get paid for it or read between the patients’ [visits]…’* (Interview 15)

#### Challenges

Lack of social interaction with other colleagues, being dependent on the Internet and technology, as well as patients’ high expectations and misconceptions about video consultations were among the problems that participants mentioned.

##### Social interaction with colleagues

Some of the participants explained that they needed to communicate with patients and colleagues in a different way in the digital care setting than a physical setting.

*‘You don’t get the social contact from your fellow colleagues as you get when you are at work [in a physical setting].’* (Interview 5)

##### Technical problems

Participants asserted some challenges and limitations regarding working in the digital care setting. The infrastructure of the communication channel in the digital care setting relies on technology and the Internet. In case of a technical problem or any interruption of the Internet, communication with the patient will be disrupted or disconnected.

*‘Since we are dependent on the internet and IT, it needs to work, some days it can be too much on the internet and it gets slower and sometimes it breaks down, but they [the technical team] are really quick. So, it is not a problem.’* (Interview 6)

*‘Obviously one thing, and that can be technical issues because it is digital. If something technically goes wrong like connection or something in the portal, I cannot do anything about it and that can be annoying if I have a meeting with the patient. I have to trust the technical team to resolve it.’* (Interview 14)

##### Patients’ high expectations

Some participants believed that in video consultations patients have unrealistic expectations and get disappointed when they do not receive care. Even though it is possible to provide care for several problems digitally, there are still some limitations which make it impossible to help all patients.

*‘Sometimes patients have no realistic expectation; they think we can make judgment of their back or knee or ear and then when we say we cannot help you with this they get a little bit disappointed. You cannot do all things but for many kinds of things digital is really good*…*’* (Interview 11)

*‘You need to examine [the patient] physically and those things you can’t do, and some patients don’t understand that.’ (*Interview 4)

##### Unfamiliarity with the culture of digital care

Another problem that nurses face is the lack of sufficient knowledge of patients about how to communicate and behave in the video consultation meetings. The nurses stated that this could affect the quality of their services.

*‘Maybe sometimes people are calling from a car or dark room. It was loud and difficult to see, like a wound on a hand, otherwise you need to hear what they are saying.’* (Interview 12)

*‘*… *Sometimes they are sitting in a car, or a dark room and I have to say I need to see you. That is a problem sometimes. Or parents calling while their kids are sleeping, and they get angry if we ask them to wake up the child.’* (Interview 11)

##### Lack of physical contact with patient

Due to the lack of physical contact with patients, participants need to use their experiences as well as different methods to obtain accurate information from the patients to be able to evaluate their needs.

*‘[In a video consultation] you are very much on your own. I use my personality to make the patient comfortable and calm. The thing you have to do is you have to be very clear with your questions, and repeat their answers, and make sure that information they are giving you is correct.’* (Interview 5)

*‘You have to think of another way because then you can ask them “can you check your pulse?” Then I look at the breath. So, you have to think in another way sometimes, but you do learn that quite quickly.’* (Interview 3)

Participants mentioned that they did not see any difference between what they could see in a face-to-face meeting and what they could see in a video consultation, and that they could make a good assessment of the patient even without touching them. The important thing is to make sure that the information they receive is precise with clear questions and answers. What participants can evaluate or do is limited to what they see and hear. If the disease requires further evaluation, such as a physical examination, the patient should be referred to a healthcare center. The participants believed that video consultation could complement physical meetings.

*‘There is not any difference between what I can see on the computer or what I can see in real life. If you have an irritation or something.’* (Interview 4)

*‘You assess them more than you think you do. You assess them without touching them. The thing you need to do is you have to be very clear with your questions, and repeat their answers, and make sure that information they are giving you is correct. You assess them like they are in the health clinic. You look at them, how they walk, how they talk.’* (Interview 5)

#### Combining digital and physical care

##### Competence development for care professionals

The participants expressed that in today’s health care, providing care digitally is a necessity. A combination of physical and digital care was appreciated by the nurses in this study.

*‘I think you cannot hold the digital meeting away. You are on the point that you need to explore the digital meeting. You need to make a combination of physical care and digital care. We have our physical clinic in Malmö, Lund and Jönköping.’ (Interview 4)*

Some of the participants were concerned about their competence development in the digital setting and expressed that it is difficult to practice the skills if they only work digitally.

*‘*… *If you do digitally all the time it would be hard. I think in a long time it will be boring with digital and when you are educated you want to do some practice. with digital you cannot practice your skills.’ (Interview 13)*

##### Benefits for patients

The Participants believed that combining the digital and physical care would be an optimal solution for providing care to patients.

*‘I would say the best for the patient and nurse is to have physical and digital. And all the time with video consultation is hard and if you can combine it is good. And also, it is good that patients can meet the nurse or the doctor physically*…*’ (Interview 13)*

## Discussion

The results of this study show that despite the limitations in digital care settings, nurses are satisfied with the use of video consultations. Flexibility, efficiency, and access to information have been some of the benefits mentioned by the participants in this study. This is also in line with previous studies in which care professionals such as physicians are satisfied and positive towards using video consultations [44–46].

The aim of this study was to explore nurses’ experience of using video consultation and its impact on their workflow and communication in Sweden and give insight into its opportunities, challenges, and limitations.

Eight categories and several subcategories were identified (Table 1). The results showed that the use of video consultations in a digital setting had a positive impact on communication between nurses, patients, and the nurses’ workflow. This is aligned with the results of the use of video consultations in a physical setting [30,31,33]. The results also showed that having access to prior knowledge about the patient, not commuting to work and no physical preparation routines enhance the efficiency of the nurses’ work process. The nurses needed mainly to focus on the patient and be fully prepared to contact him/her. Although nurses are satisfied with the opportunity to work from home, having autonomy about choosing their working hours, there are still some challenges that need to be considered. Some of these challenges include lack of social interaction with co-workers, being dependent on technology, fast changes of the platform, facing patients’ unrealistic expectations about digital healthcare services, dealing with patients’ unfamiliarity with the culture of digital care, and lack of physical contact with patients. These challenges have also been mentioned in some previous studies [10,44,45,46]. Furthermore, the nurses in this study as participants in some other studies felt that technical problems may affect their workflow [32,45]. Improvement of technical solutions has been seen as a necessity in a study focusing on the challenges of video consultations [13].

Findings of this study showed that nurse-patient communication during video consultation was relevantly comparable with the physical meetings. The participants mentioned that they could focus on patients during the video consultations and not get disrupted by other factors. Hence, the findings of this study do not confirm the results of the previous study on the nurse’s perception regarding nurse-patient communication while providing care via telehealth [47]. That study revealed that telehealth facilitates nurses work process but impedes the communication process due to struggling to understand non-verbal signals [47]. In addition, patient expectations need to be balanced and digital consultations culture needs to be accepted and respected by the patients as high expectations and unfamiliarity with the digital setting may result in difficulties in provision of online care [13].

Nurses’ communication with each other was faster and easier as they used different communication channels and had access to the conversation history. However, the results showed that communication and providing diagnosis were easier for experienced nurses as they could use their previous communication skills to establish a relationship with the patients and provide a diagnosis despite the limitations with digital care. This result also has been confirmed in a previous study[47]. Since the importance of communication has been mentioned in previous studies, having a training program in which experienced nurses could help junior nurses in communication with the patients and other team members in a digital care setting could be beneficial [47].

The results in this study, showed that the patient got referred to a medical center if a physical examination was needed. The nurses mentioned that providing health care only digitally will not cover all aspects needed to treat a patient or all types of diseases.

There is, therefore, a need of combining digital with physical care in cases in which only digital care is not the optimal way to provide health care.

The video consultations, due to covid-19 situation and the current restrictions, can increase cooperation with physical healthcare centers. However, video consultations cannot be a substitute for face-to-face visits but can complement them and reduce the need of face-to-face consultations in some situations e.g., during the COVID-19 pandemic. This is in line with the results of a scoping review regarding clinicians’ experience of using video consultations in primary care [48]. However, some patients prefer face-to-face consultation when it is possible [10].

In this study we provided further insight into the digital setting. There have been studies exploring care professionals’ experience of using video consultations, however, there was a lack of studies focusing on nurses’ workflow and communication in the digital care settings and our study contributes to this knowledge gap. However, more studies are needed within this area as we in this study only interviewed nurses working for one online private care provider. The results may differ from other healthcare providers using different digital platforms. In addition, further studies can be conducted not only in private sectors but also in public sectors and even considering other aspects of the socio-technical model such as internal organizational features, external rules, and regulations and their impact on nurses’ workflow in a digital setting.

## Conclusion

Even though providing online care has its limitations, the nurses were positive towards using video consultations. The opportunities such as flexibility, efficiency in the nurses’ workflow and information accessibility have been perceived as strengths in digital settings.

The occurrence of the COVID-19 pandemic during two previous years has caused an increasing use of video consultations worldwide. This fundamental change requires extensive research within this area from both patients, their next-of-kin and care professionals’ perspective. In addition, the COVID-19 pandemic brought the realization that providing digital healthcare meetings to patients and citizens is a necessity in future health care.

## Data Availability

The datasets generated during the study are not publicly available. Sharing the qualitative data collected through the interviews in this study may jeopardize the study participants privacy.

## Acknowledgment

We would like to thank all the study participants for their time and valuable insights.

